# Previous COVID-19 infection and antibody levels after vaccination

**DOI:** 10.1101/2021.09.04.21263121

**Authors:** Hamad Ali, Barrak Alahmad, Abdullah A. Al-Shammari, Abdelmohsen Al-Terki, Maha Hammad, Preethi Cherian, Irina Alkhairi, Sardar Sindhu, Thangavel Alphonse Thanaraj, Anwar Mohammed, Ghazi Alghanim, Sriraman Deverajan, Rasheed Ahmad, Sherief El-Shazly, Ali A. Dashti, Mohammad Shehab, Salman Al-Sabah, Abdullah Alkandari, Jehad Abubaker, Mohamed Abu-Farha, Fahd Al-Mulla

## Abstract

**Background:** The emergence of new COVID-19 variants of concern coupled with a global inequity in vaccine access and distribution, prompted many public health authorities to circumvent the vaccine shortages by altering vaccination protocols and prioritizing high-risk individuals. Those with previous COVID-19 infection may have not been prioritized due to existing humoral immunity.

**Objective:** We aim to study the association between previous COVID-19 infection and antibody levels after COVID-19 vaccination.

**Methods:** A serological analysis to measure SARS-CoV-2 IgG, IgA and neutralizing antibodies was performed on individuals who received one or two doses of either BNT162b2 or ChAdOx1 vaccines in Kuwait. Generalized linear regression models adjusted for individual characteristics and comorbidities were fitted to study the average levels of IgG and neutralizing antibodies in vaccinated individuals based who had previous COVID-19 infection compared to those who had not.

**Results:** A total of 1025 individuals were recruited. The mean levels of IgG, IgA and neutralizing antibodies were higher in vaccinated subjects with previous COVID-19 infection when compared with those vaccinated without previous COVID-19 infection. Regression analysis showed a steeper slope of decline for IgG in vaccinated individuals without previous COVID-19 infection in comparison with vaccinated individuals with previous COVID-19 infection.

**Conclusion:** Previous COVID-19 infection appears to elicit robust and sustained levels of SARS-CoV-2 antibodies in vaccinated individuals. Given the inconsistent supply of COVID-19 vaccines in many countries due to the global inequity, our results point towards wider vaccination plans to especially cover individuals without previous COVID-19 infection.

## Introduction

The COVID-19 pandemic continues to affect global health in an unprecedented scale with more than 200 million cases and claiming more than 4 million lives worldwide to date, with ongoing strains on the global economy (Dong et al., 2020). Non-pharmaceutical interventions were implemented worldwide since the outbreak started in attempts to slow down the pandemic waves while efforts have been focused on the development of effective COVID-19 vaccines to curtail the pandemic especially among vulnerable subpopulations (Al-Shammari et al., 2020; Khadadah et al., 2021).

The World Health Organization (WHO) and the US Food and Drug Administration (FDA) had declared the release of COVID-19 vaccines in September 2020 (Kaur and Gupta, 2020). Between December 2020 and February 2021, adenoviral vector vaccines like ChAdOx1 (AstraZeneca-Oxford) and mRNA vaccines like BNT162b2 (Pfizer BioNTech) and mRNA-1273 (Moderna) were put in use. The three vaccines; ChAdOx1, which consists of non-replicative simian adenovirus vector with the full-length code of the spike protein of severe acute respiratory syndrome coronavirus 2 (SARS-CoV-2) and the other two which utilize a novel mRNA vaccine platform, have shown acceptable safety and efficacy profiles in clinical trials (Folegatti et al., 2020; Polack et al., 2020; Baden et al., 2021). Remarkably, these vaccines, which are administrated in two intramuscular shots regimes, have shown to offer protection by eliciting the production of anti-S-RBD (S-protein receptor binding domain) immunoglobulin G (IgG), IgM, and IgA isotypes, with neutralization activity capable of inhibiting the RBD binding to ACE2 cognate receptor (Sahin et al., 2020; Ewer et al., 2021). The measure of these antibodies in sera and, in particular, the neutralizing antibody levels can provide an indication of the level of protection induced by either COVID-19 vaccination or previous infection (Khoury et al., 2021).

There is a great argument concerning the nature, stability, and durability of antibody responses over time in COVID-19 patients, with several studies reporting stable antibody persistent immunity (Gudbjartsson et al., 2020) and others showing rapid antibody waning immunity, or late appearance with low antibody levels and/or complete lack of long-lasting antibodies (Jeyanathan et al., 2020; Ali et al., 2021). Further studies are needed to elucidate the safety and efficacy of a booster, especially in immunocompromised and/or immunosuppressed individuals in order to determine the best dosing schedule and mix-and-match schedules of vaccines. The US public health officials urged FDA to decide on booster vaccines as the benefits of COVID-19 vaccination far outweigh the potential risks (Tanne, 2021). Several countries have announced plans for booster-shot programs (Callaway, 2021). Third doses of vaccines developed by Moderna, Pfizer–BioNTech, Oxford–AstraZeneca and Sinovac prompted a spike in levels of infection-blocking ‘neutralizing’ antibodies, when administered several months after the second dose (Callaway, 2021).

On the other hand, previously infected individuals were shown to have some humoral protection against COVID-19 (Abbasi, 2021), although still prone to reinfection (Stokel-Walker, 2021; Wang et al., 2021). Meanwhile, calls for global vaccine equity continue to fail as many developing countries are incapable of obtaining enough vaccines to protect a substantial proportion of the population. Towards this end, many countries attempted to ration their vaccine supplies by prioritizing individuals that were not previously infected. In Kuwait, those who had previous COVID-19 infection were temporarily given a single dose of vaccine while those who did not get the infection were prioritized to receive two doses. It is of interest to investigate the levels of humoral antibody response comparing those who were previously infected to those who were not.

This study specifically assessed the antibody-mediated immune response in terms of SARS-CoV-2 S1 specific IgG, IgA, IgM and anti-S RBD neutralizing antibody levels produced in vaccinated people who were either previously infected with COVID-19 or did not get infected in Kuwait.

## Material and Methods

### Participant’s recruitment

The Ethical Review Committee of Dasman Diabetes Institute and Kuwait Ministry of Health ethical committee reviewed and approved the study as per the updated guidelines of the Declaration of Helsinki and of the US Federal Policy for the Protection of Human Subjects (approval references; RA HM-2021-008 for ERC and 3799). The study recruited people who received either one or two doses of BNT162b2 (Pfizer–BioNTech) mRNA vaccine and ChAdOx1 (AstraZeneca) vaccine. The two groups were further divided into subgroups based on previous COVID-19 infection. The diagnosis of COVID-19 was established based on a positive SARS-CoV2 PCR result for a nasopharyngeal swab. Participants taking immunosuppressants and/or chemotherapy were excluded from the study. RedCap survey was used to collect data from each participant including age, gender, chronic health conditions, height, weight, and self-reported history of COVID-19 infection. Samples were collected at Dasman Diabetes Institute after signed informed consent was obtained from all participants.

### Blood sample collection

Blood samples were obtained from participants and collected in EDTA tubes. Samples were centrifuged at 400xg for 10 minutes at normal room temperature to separate the plasma which was then aliquoted and stored at −80°C.

### Quantification of plasma levels of SARS-CoV-2-specific IgG, IgM and IgA

Enzyme-linked immunosorbent assay (ELISA) kit (SERION ELISA agile SARS-CoV-2 IgG and IgA SERION Diagnostics, Würzburg, Germany) were used to determine plasma levels of SARS-CoV-2-specific IgG and IgA antibodies following the manufacturers protocol. The positive and negative cutoffs were determined as per manufacturer recommendations. The IgG levels were reported as binding antibody units (BAU)/mL. values of < 21 BAU/mL were considered negative where values of 21.0-31.5 BAU/mL were considered as border line and levels higher than 31.5 BAU/mL were considered as positive. The IgM levels were reported as Arbitrary Units (AU)/mL. values of < 90 AU/mL were considered negative where values of 90-110 AU/mL were considered as border line and levels higher than 110 AU/mL were considered as positive. The IgA levels were reported as Arbitrary Units (AU)/mL. values of < 10 AU/mL were considered negative where values of 10-14 AU/mL were considered as border line and levels higher than 14 AU/mL were considered as positive.

### Quantification of plasma levels of SARS-Cov-2-specific Neutralizing Antibodies

SARS-CoV-2-specific surrogate Virus Neutralization Test (sVNT) was utilized to study the levels of plasma neutralizing antibodies against SARS-CoV-2 S-RBD (SARS-Cov-2 sVNT kit, GenScript, USA, Inc). We followed manufacturers protocol. Evaluation of resulted data was performed following manufacturers recommendations for positive and negative cutoffs. Results were construed by calculating inhibition rates for samples as per the following equation: Inhibition = (1 - O.D. value of sample/O.D. value of negative control) ×100%. Neutralizing antibody levels higher than 20% were considered positive.

### Statistical analysis

The descriptive data was summarized using mean, median, standard deviation and interquartile range where applicable. For the regression analysis, we restricted the data to those who have received both doses of BNT162b2. We built generalized additive linear models where plasma IgG levels was fitted as the dependent variable adjusting for type-2 diabetes status (yes/no), hypertension (yes/no), BMI (linear), age (smoothed spline), gender, comorbidity score (sum score of equal weight for heart disease, stroke, chronic obstructive pulmonary disease, asthma, obstructive sleep apnea, chronic kidney disease, bleeding disorders and other chronic diseases), previous COVID-19 infection (yes/no) and duration since receiving the last vaccine dose (smoothed spline) as the independent variables. Penalized splines were fitted for two continuous variables; age and duration since last dose to explore and control for non-linearity using restricted maximum likelihood (REML) estimation. The smoothing penalized splines are nonparametric terms which can optimize the goodness-of-fit by cross-validation through a penalty term for over- and under-fitting. We determined and interpreted the adjusted effect estimate with 95% confidence intervals as changes in the mean IgG levels comparing; 1) those with previous infection to those who did not get an infection; and 2) the estimated decline for every 100 days since receiving the second dose. We further studied the interaction analyses looking into the effects of having previous infection across the decline in IgG levels since receiving the second dose of the vaccine. on the slope of IgG decline over time. We included an interaction term between previous COVID-19 infection status and the duration (in days) since receiving the second to examine effect measure modification and reported the Wald-test p-value for statistical significance. All analyses were performed using R software version 3.3.1 (R Foundation for Statistical Computing), penalized splines were implemented in generalized additive models using the *mgcv* package. A p-value of less than 0.05 was considered to indicate statistical significance.

## Results

### Cohort characteristics

The study included 1025 participants who were descriptively divided into 4 groups based on the number of doses received and the type of vaccine. 41 subjects received one dose of BNT162b2, 490 subjects received two doses of BNT162b2, 299 subjects received one dose of ChAdOx1 and 195 subjects received two doses of ChAdOx1. Each group was subdivided into two subgroups based on previous infection status. The distribution of the participants and their age in each group is presented in Table.1 and Table.2.

**Table 1.**
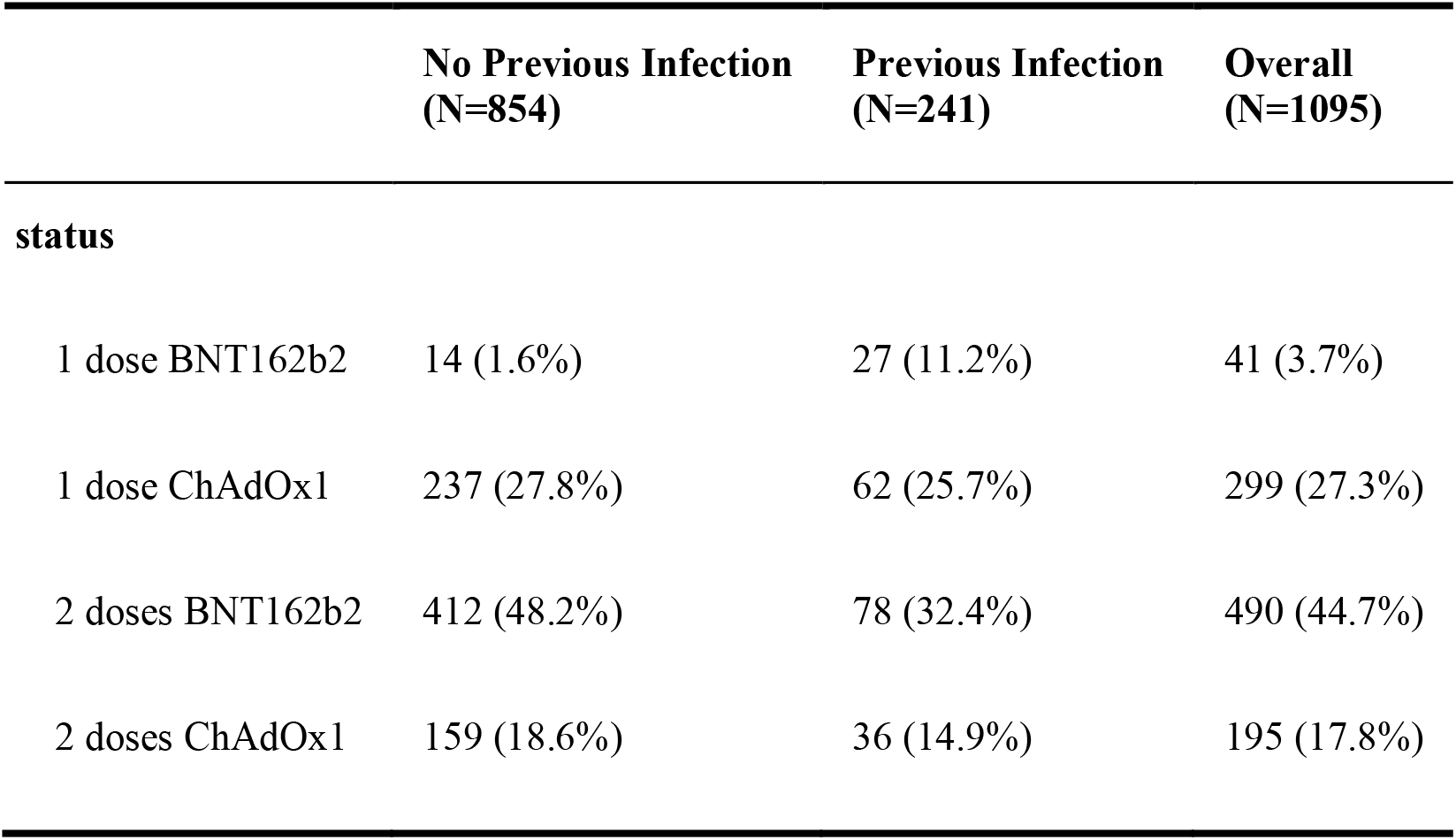
Cohort vaccination status stratified by previous COVID-19 infection.

**Table 2.** Cohort characteristics and serological results.

|  | 1 dose BNT162b2 |  | 1 dose ChAdOx1 |  | 2 doses BNT162b2 |  | 2 doses ChAdOx1 |  | Overall |  |
| --- | --- | --- | --- | --- | --- | --- | --- | --- | --- | --- |
|  | No Previous Infection<br>(N=14) | Previous Infection<br>(N=27) | No Previous Infection<br>(N=237) | Previous Infection<br>(N=62) | No Previous Infection<br>(N=412) | Previous Infection<br>(N=78) | No Previous Infection<br>(N=159) | Previous Infection<br>(N=36) | No Previous Infection<br>(N=822) | Previous Infection<br>(N=203) |
| **Age (Years)** |  |  |  |  |  |  |  |  |  |  |
| Mean (SD) | 47.6<br>(12.6) | 42.3<br>(11.6) | 48.3<br>(11.0) | 46.3<br>(9.24) | 48.1<br>(14.2) | 46.0<br>(13.8) | 43.1<br>(11.4) | 40.4<br>(10.3) | 47.2<br>(12.9) | 44.6<br>(11.8) |
| Median<br>[Min,<br>Max] | 44.3<br>[21.5,<br>66.0] | 42.4<br>[22.5,<br>73.2] | 51.1<br>[21.0,<br>74.0] | 47.5<br>[24.3,<br>63.6] | 48.3<br>[21.8,<br>87.4] | 42.7<br>[21.0,<br>74.2] | 42.5<br>[21.0,<br>89.1] | 38.0<br>[21.5,<br>65.0] | 47.6<br>[21.0,<br>89.1] | 42.9<br>[21.0,<br>74.2] |
| **Duration (Days)** |  |  |  |  |  |  |  |  |  |  |
| Mean (SD) | 31.8 | 56.7 | 91.6 | 99.4 | 91.7 | 82.1 | 30.0 | 28.9 | 78.7 | 74.6 |
|  | (19.3) | (31.2) | (25.6) | (27.1) | (45.1) | (45.4) | (12.4) | (11.9) | (43.3) | (42.2) |
| Median<br>[Min,<br>Max] | 22.0<br>[7.00,<br>76.0] | 52.0<br>[9.00,<br>124] | 99.0<br>[19.0,<br>150] | 105<br>[21.0,<br>159] | 90.0<br>[6.00,<br>197] | 76.0<br>[12.0,<br>195] | 26.0<br>[1.00,<br>61.0] | 27.0<br>[5.00,<br>48.0] | 78.0<br>[1.00,<br>197] | 72.0<br>[5.00,<br>195] |
| **IgG (BAU/ml)** |  |  |  |  |  |  |  |  |  |  |
| Mean (SD) | 157<br>(63.5) | 195<br>(39.9) | 80.0<br>(70.1) | 155<br>(61.2) | 137<br>(55.1) | 188<br>(42.7) | 116<br>(50.5) | 146<br>(42.2) | 117<br>(64.1) | 172<br>(52.1) |
| Median<br>[Min,<br>Max] | 172<br>[5.10,<br>226] | 204<br>[67.6,<br>263] | 51.6<br>[1.42,<br>261] | 177<br>[5.85,<br>230] | 141<br>[11.0,<br>260] | 198<br>[35.5,<br>266] | 116 [0,<br>245] | 155<br>[5.94,<br>209] | 122 [0,<br>261] | 185<br>[5.85,<br>266] |
| Missing | 0 (0%) | 0 (0%) | 0 (0%) | 1 (1.6%) | 0 (0%) | 0 (0%) | 0 (0%) | 0 (0%) | 0 (0%) | 1 (0.5%) |
| **Neutralizing Antibodies (%)** |  |  |  |  |  |  |  |  |  |  |
| Mean (SD) | 76.9<br>(25.8) | 90.8<br>(7.14) | 49.6<br>(35.9) | 83.1<br>(24.1) | 82.2<br>(16.1) | 91.0<br>(10.7) | 82.8<br>(18.7) | 90.6<br>(12.8) | 72.7<br>(28.4) | 88.4<br>(16.5) |
| Median<br>[Min,<br>Max] | 86.8<br>[3.70,<br>94.3] | 93.5<br>[65.5,<br>95.3] | 38.7 [0,<br>95.1] | 93.7<br>[10.3,<br>95.3] | 88.7 [0,<br>95.1] | 94.1<br>[28.2,<br>95.8] | 90.4<br>[3.92,<br>95.1] | 93.4<br>[22.4,<br>95.1] | 87.2 [0,<br>95.1] | 93.7<br>[10.3,<br>95.8] |
| Missing | 0 (0%) | 1 (3.7%) | 2 (0.8%) | 1 (1.6%) | 17<br>(4.1%) | 8<br>(10.3%) | 3 (1.9%) | 3 (8.3%) | 22<br>(2.7%) | 13<br>(6.4%) |
| **IgM (AU/ml)** |  |  |  |  |  |  |  |  |  |  |
| Mean (SD) | 141<br>(201) | 126<br>(123) | 39.7<br>(70.0) | 58.6<br>(78.9) | 50.7<br>(76.0) | 79.6<br>(88.4) | 32.5<br>(67.5) | 42.3<br>(50.7) | 45.5<br>(77.7) | 72.8<br>(88.8) |
| Median<br>[Min,<br>Max] | 71.9<br>[5.50,<br>800] | 81.7<br>[6.70,<br>407] | 13.3 [0,<br>494] | 30.0<br>[0.800,<br>474] | 23.6 [0,<br>600] | 50.4<br>[6.50,<br>600] | 15.7<br>[1.43,<br>551] | 24.2<br>[5.07,<br>250] | 18.8 [0,<br>800] | 40.4<br>[0.800,<br>600] |
| **IgA (AU/ml)** |  |  |  |  |  |  |  |  |  |  |
| Mean (SD) | 30.5<br>(29.2) | 66.1<br>(40.9) | 11.5<br>(22.5) | 41.6<br>(42.9) | 22.8<br>(34.5) | 69.2<br>(48.3) | 11.6<br>(20.9) | 31.9<br>(26.1) | 17.5<br>(29.5) | 53.8<br>(44.8) |
| Median<br>[Min,<br>Max] | 19.6<br>[3.34,<br>102] | 55.7<br>[14.3,<br>155] | 2.78<br>[0.0400,<br>200] | 27.0<br>[1.19,<br>200] | 9.42 [0,<br>200] | 58.3 [0,<br>200] | 4.17<br>[0.151,<br>130] | 25.8<br>[0.422,<br>93.9] | 6.29 [0,<br>200] | 38.1 [0,<br>200] |
| Missing | 2<br>(14.3%) | 3<br>(11.1%) | 34<br>(14.3%) | 12<br>(19.4%) | 38<br>(9.2%) | 8<br>(10.3%) | 2 (1.3%) | 1 (2.8%) | 76<br>(9.2%) | 24<br>(11.8%) |

### Levels of antibodies according to previous infection status

We descriptively compared mean levels of SARS-CoV-2 IgG, IgM, IgA and Neutralizing Antibodies in the groups based on COVID-19 previous infection status. In each group, the mean levels of SARS-CoV-2 IgG, IgA and Neutralizing Antibodies were higher in subgroups of vaccinated subjects with previous COVID-19 infection when compared with those vaccinated without previous COVID-19 infection (Table.2) and (Figure.1–2). The mean duration between the serology test and last dose of vaccine taken was 74.6 ± 42.3 days for subjects with previous infection and 78.7 ± 43.3 days for those without previous infection (Table.2). Overall, the mean level of IgG in vaccinated individuals with previous COVID-19 infection who received one or two doses of either vaccines was 172 ± 52.1 BAU/ml in comparison to 117 ± 64.1 BAU/ml in those without previous COVID-19 infection (Table.2). The mean level of IgA in vaccinated individuals with previous COVID-19 infection who received one or two doses of either vaccines was 53.8 ±44.8 AU/ml in comparison to 17.5 ±29.5 AU/ml in those without previous COVID-19 infection (Table.2). While the mean percentage of Neutralizing Antibodies in vaccinated individuals with previous COVID-19 infection who received one or two doses of either vaccines was 88.4 ± 16.5 % in comparison to 72.7 ± 28.4 % in those without previous COVID-19 infection (Table.2).

**Figure 1.**
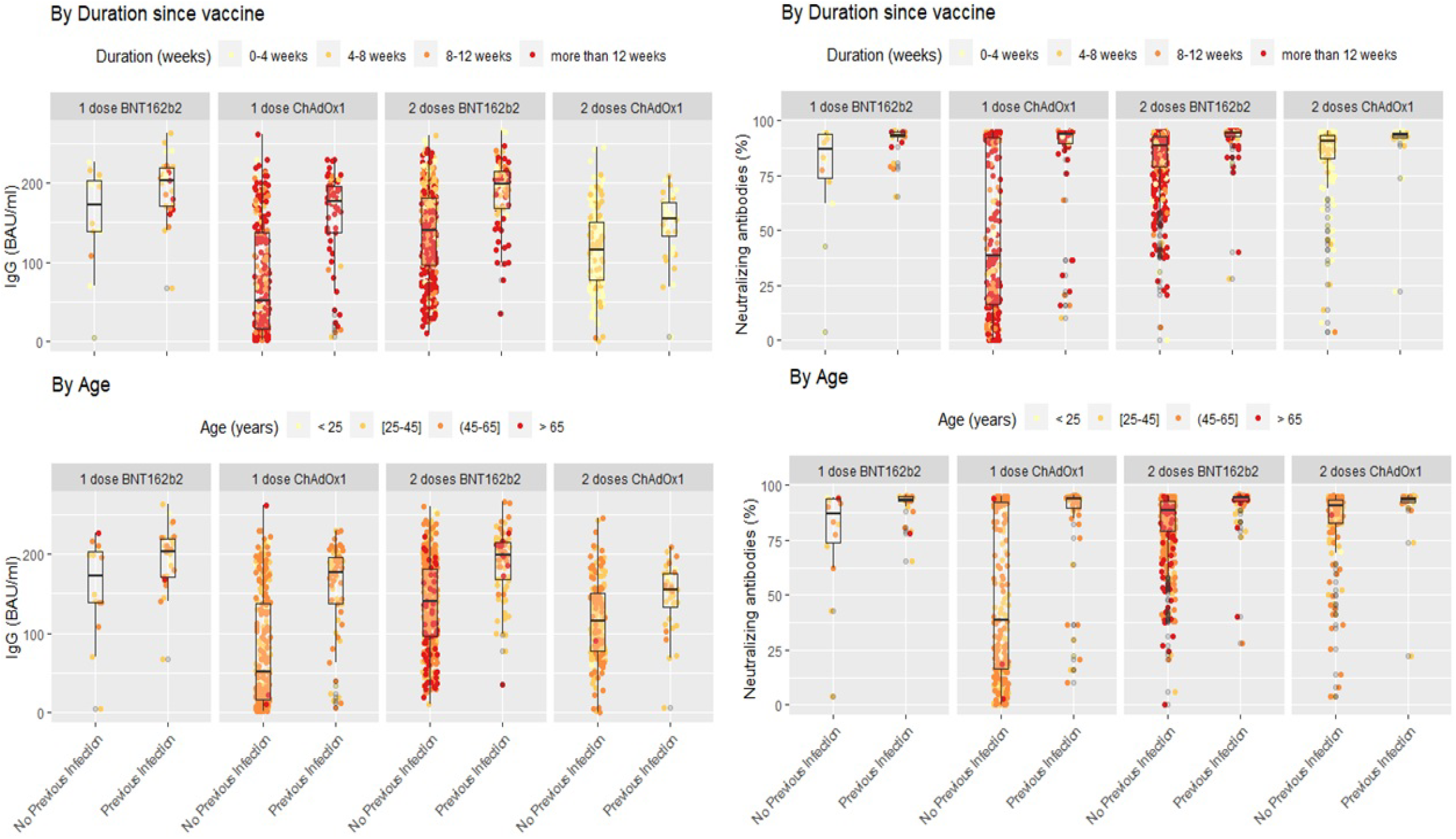
SARS-COV2 IgG and neutralizing antibodies in vaccinated individuals stratified by previous infection status while duration since vaccination and age variations shown based on color coding. In each subgroup individuals with previous COVID-19 infection showed higher levels of IgG and Neutralizing antibodies in comparison with their counterparts who did not encounter COVID-19. Lower levels of IgG seem to be associated with a cluster of older individuals who received two doses of BNT162b2 without previous infection.

**Figure 2.**
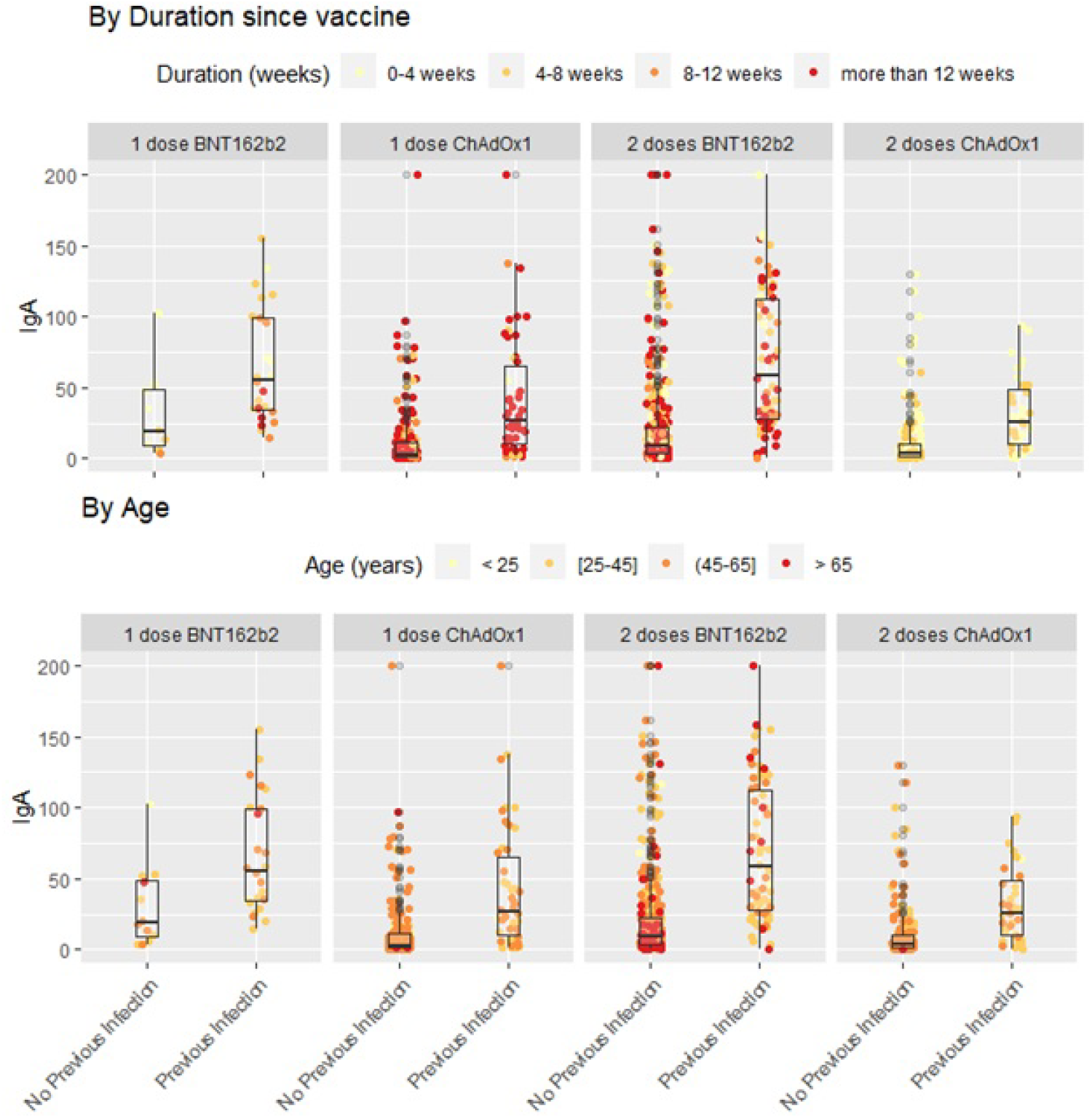
SARS-COV2 IgA in vaccinated individuals stratified by previous infection status while duration since vaccination and age variations shown based on color coding. In each subgroup individuals with previous COVID-19 infection showed higher levels of IgA in comparison with their counterparts who did not encounter COVID-19. Lower levels of IgA seem to be associated with a cluster of older individuals who received two doses of BNT162b2 without previous infection.

### Regression Analyses

Generally, IgG declined linearly as more days passed after receiving the vaccine dose. Average IgG levels declined by −60.05 BAU/mL (95% CI; −68.98, −51.11; p<0.001) for every 100 days passed since the second vaccine (Table 3). IgG-duration adjusted smooth relationships with 95% confidence intervals is presented in Figure 3. Among vaccinated individuals, those with previous COVID-19 infection had, on average, higher IgG levels by 46.65 BAU/mL (95% CI; 35.85, 57.45; p<0.001).

**Table 3.** Change in IgG levels (in BAU/ml) in adjusted multiple linear models for those who received 2 doses of BNT162b2.

| Variable | Change | 95%<br>low | 95%<br>high | p-value |
| --- | --- | --- | --- | --- |
| **Overall** |  |  |  |  |
| Previously infected vs. No previous infection | 46.65 | 35.85 | 57.45 | <0.001 |
| Decline for every 100 days | -60.05 | -68.98 | -51.11 | <0.001 |
| **Interaction** |  |  |  |  |
| Decline for every 100 days & no previous infection | -63.40 | -73.09 | -53.71 | p-value for interaction = 0.084 |
| Decline for every 100 days & previously infected | -42.28 | -64.31 | -20.25 |  |

**Figure 3.**
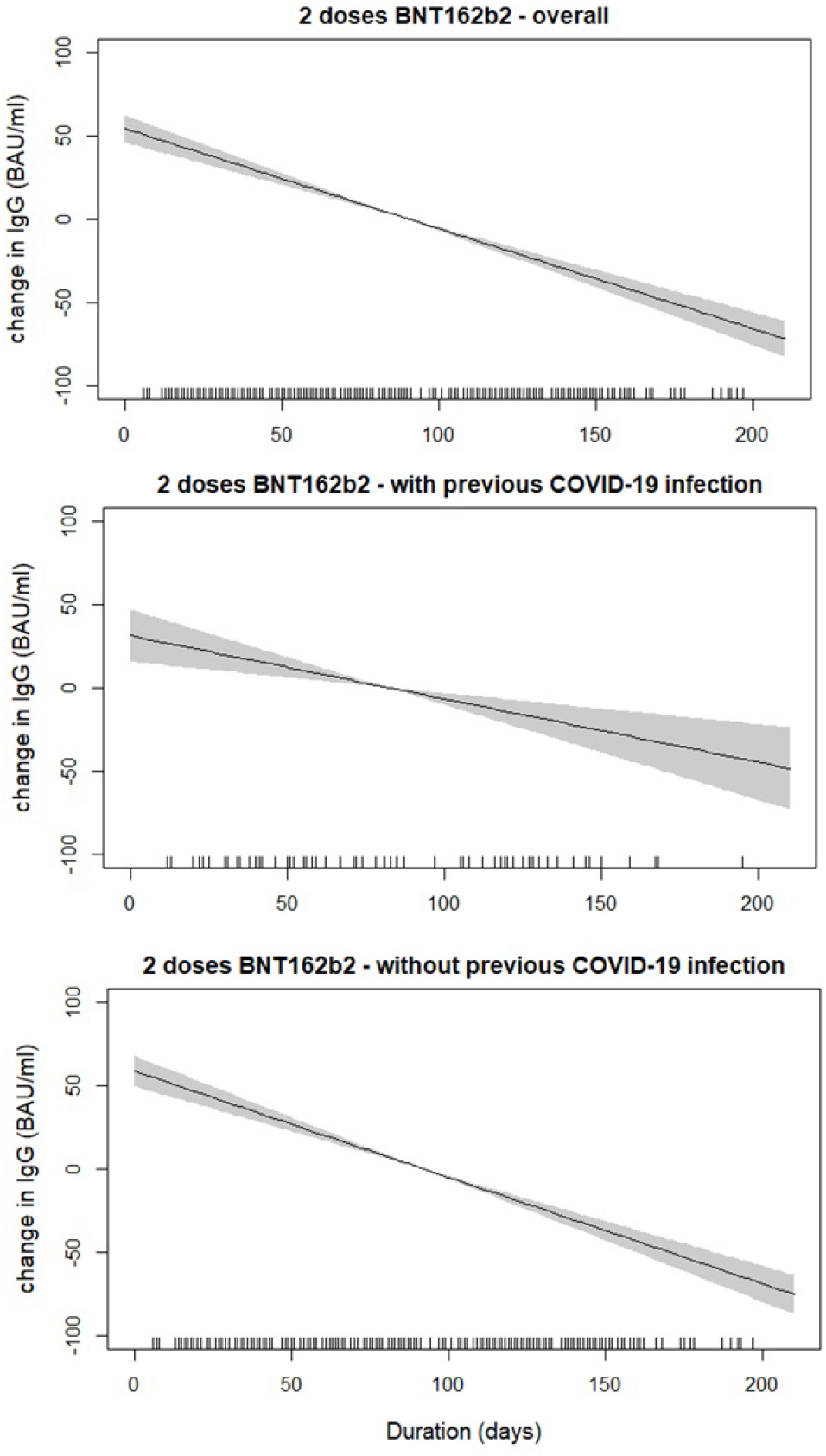
SARS-COV2 IgG antibodies decline over time since receiving the second dose of BNT162b2 (Pfizer–BioNTech) vaccine stratified by previous COVID-19 infection status. The smooth relationships were derived from generalized additive models with penalized splines for duration (in days) and adjusted for age, gender, BMI and comorbidity scores. Solid lines represent the effect estimates of change over time while the shaded areas represent the 95% confidence intervals. The graphs show faster decline for both markers in vaccinated individuals without previous COVID-19 infection in comparison to those with previous infection.

When the linear decline in subjects receiving 2 doses of BNT162b2 was compared between those with previous COVID-19 infection versus those without, we found that the IgG slope of the group without infection was steeper (Table.3). In the interaction analyses, the group with 2 doses of BNT162b2 without previous infection was associated with lower IgG levels as compared to those with previous infection (p-value for interaction = 0.084) (Table.3). However, the interaction terms were likely not powered to result in statistically significant p-values.

## Discussion

Unquestionably, the best strategy to fight the COVID-19 pandemic was represented by a global vaccination campaign able to administer safe and effective vaccines. However, this was challenged by inequitable vaccine distribution and access across countries. The COVID-19 vaccination campaign started in Kuwait in 2021 with priority given to high risk groups including healthcare workers, the elderly and the immunocompromised patients with pre-existing conditions. With the COVID-19 vaccine supply chain struggles, at times, to match the increasing demands, health authorities were obliged to develop vaccination strategies to cope with inconsistent supply of COVID-19 vaccines. This included a single-dose provision to individuals who were previously infected in Kuwait. We relied on observational data from self-recruitment to measure antibody levels in Kuwait. Due to the limited data and the differential vaccine type received, we were not able to compare a single dose with infection to those with no previous infection and two doses. However, we indirectly investigated the effect of a previous COVID-19 infection on the SARS-CoV-2 antibodies among all individuals who completed two doses of vaccination. Such findings have the potential to help in vaccination prioritization strategies to cope with any shortage of vaccine supply.

The main finding of this study is that there were significantly higher levels of antibodies in fully-vaccinated individuals with previous COVID-19 infection (natural immunity) as compared to fully-vaccinated individuals without prior infection (acquired immunity). Additionally, we found that those without previous infection showed a faster decline in antibodies over time. This difference based on infection status was not surprising and could be attributed to several reasons. First, we still do not have a definite answer on how long antibodies would last after a COVID-19 infection or vaccination. Many studies are still detecting the antibodies after several months of infection regardless of the severity of the disease with one study showing detectable levels of Neutralizing antibodies seven months post-infection (Ripperger et al., 2020). Similarly, studies on vaccines are showing a strong antibody response. A clinical trial on the Moderna vaccine reported high levels of antibodies after six months of administering the second dose (Doria-Rose et al., 2021). Another study on ChAdOx1 showed high levels of antibodies after three months of a single dose (Voysey et al., 2021). Considering that the antibody-making B cells would multiply after each exposure; whether due to the infection or the vaccine, the high antibody levels in the previously infected groups are most probably representing an additive amount of the antibodies produced after the infection and after vaccination. Second, while the vaccines work by eliciting a similar immune response to a viral infection, they present the viral protein in slightly different conformations (Greaney et al., 2021). In addition, and specifically for BNT162b2, mRNA delivery using lipid nanoparticles could be presenting the antigens to the immune system in a different way than the actual viral infection and this could result in some difference in antigens kinetics and the antibodies produced (Pardi et al., 2018; Dai and Gao, 2021). Finally, natural immunity and acquired immunity can differ in the types of antibodies produced. COVID-19 vaccines expose the immune system only to a certain part of the virus. Therefore, the immune system might not be producing as many different types of antibodies after vaccination as it would after an actual COVID-19 infection. The same reasons can be used to explain the observation that a previous infection with one dose of either vaccine resulted in higher levels of antibodies compared to two doses without a previous infection.

In line with our findings, some studies are reporting similar observations in smaller cohorts. For example, a study on 51 health care workers from UK showed that previously infected individuals expressed higher levels of SARS-CoV-2 anti-S antibody titers after a single dose of BNT162b2 compared to individuals with no previous COVID-19 infection (Manisty et al., 2021). In another study that was also conducted on healthcare workers, the authors observed that participants with prior infection had antibody titers one order of magnitude higher than those without a previous infection and this was not affected by ethnicity or sex (Abu Jabal et al., 2021). In another study on BNT162b2, the authors reported similar IgG antibody levels and ACE2 antibody binding inhibition responses from a single dose in individuals with prior COVID-19 infection compared to levels measured after two doses of vaccine in individuals without prior infection (Ebinger et al., 2021). An interesting study that also reported similar conclusions to those reported here was conducted on autoimmune rheumatic diseases (AIRD) who are on immunosuppressants (Shenoy et al., 2021). The study showed that one dose of ChAdOx1 with a previous infection was also superior compared to two doses in this subset of immunocompromised patients. Altogether, data from the present study combined with findings from previous work suggest that COVID-19 infection results in a broader immune response.

The decline in IgG levels over time was expected since this is the case for all other vaccinations. The question remains on the maximum duration these antibodies would last. The serological analysis in this study was performed after an average of less than three months of vaccination. Therefore, it just confirms previous findings that showed the antibodies to last for several months after vaccination. Additionally, we observed a faster slope of linear decline of IgG levels over time among those who were not previously infected compared to those who were infected. While this can be explained by the assumed additive amount of antibodies produced after the infection and after vaccination, this phenomenon should rather be investigated in a follow-up longitudinal epidemiological design where the two groups can be fairly compared with little concerns about time-invariant confounders such as age, sex, BMI, etc.

Although we are optimistic that the ongoing vaccination should provide some sort of control on the spread of the disease, herd immunity might be difficult to achieve as current vaccines will not eliminate the infection but reduce the degree of morbidities and mortalities. Therefore, these vaccines will likely be seasonal and proper protocols need to be placed. In case of limited supply, the immunocompromised people and individuals who weren’t previously infected may need to be covered more promptly as they may still be susceptible for infection after vaccination. Overall, the guidance for urgent need of booster shots requires continuous monitoring of the antibody levels in vaccinated individuals to maintain protective COVID-19 immune response.

The emergence of the Delta variant (B.1.617), and its fast spread around the globe poses a serious problem for achieving herd immunity as current vaccines appear limited in blocking the transmission of the Delta variant, the current dominant strain. This is supported by several reports highlighting breakthrough infections of Delta in fully vaccinated individuals (Brown et al., 2021; Tang et al., 2021). As the economic, political and psychological burden of the pandemic has been significant, countries around the globe are opening up and public health measures are being relaxed and disengaged. This might allow the Delta variant to circulate more effectively resulting in new global COVID-19 waves, paving the way for a new approach to, perhaps, achieve herd immunity through what can be described as hybrid immunity. On one hand, the current vaccines provide effective protection against severe COVID-19 infections and death, viral exposure, on the other hand, might provide the needed broader immunity to block further transmission and hence, in principle, provide the lacking factor to surge towards herd immunity.

This study has a number of limitations. First, we only descriptively compared the different sub-stratification by vaccine type, number of doses and previous infection. The data was not collected to investigate such question and hence it lacked the statistical power. Secondly, the regression analyses were adjusted for a number of *a priori* confounders, however, we cannot rule out potential residual confounding by other variables such as severity of chronic illness or duration since previous infection. Finally, the cross-sectional data are prone to confounding by time-invariant such as individual characteristics. Longitudinal analyses of both humoral and cellular responses with longer follow-ups are warranted to carefully assess the duration of immunity after vaccination.

## Conclusions

In conclusion, the findings from this study confirms that prior infection with COVID-19 combined with vaccination results in a stronger antibody response when compared to vaccination without prior infections. These findings can help in implementing strategies for vaccination policies under the current inequitable distribution and access of vaccines especially when deciding on the individuals who should be prioritized.

## Data Availability

Data will be available upon request

## Authors Contribution

H.A. and M.A. Conceived and designed the analysis, researched data and wrote the manuscript. B.A. and A.A.1 performed the analysis and generated the figures. AA2, M.H.1, I.A and P.C. involved in sample collection and laboratory analysis. S.D, GA and AM involved in subjects’ recruitment and reviewed/edited the manuscript. S.A., M.S., S.E. and A.D. and AA3 involved in the clinical design of the study. S.S, T.T. and R.A. edited the manuscript. J.A. and F.A. contributed to the discussion and reviewed/edited the manuscript. All authors read and approved the manuscript.

